# Relative Effectiveness of Four Doses Compared to Three Dose of the BNT162b2 Vaccine in Israel

**DOI:** 10.1101/2022.03.24.22272835

**Authors:** Sivan Gazit, Yaki Saciuk, Galit Perez, Asaf Peretz, Virginia E. Pitzer, Tal Patalon

## Abstract

**Objectives:** The rapid spread of the Omicron variant (B.1.1.529) alongside evidence of a relatively rapid waning of the third dose prompted Israel to administer a fourth dose of the BNT162b2 vaccine on January 2022. Thus far, sufficient real-world evidence demonstrating the effectiveness of a fourth dose against infection and severe COVID-19 are lacking. This study examined the short-term effectiveness of a fourth dose compared to three doses over the span of 10 weeks.

**Design:** A retrospective test-negative case-control study, performing both a matched analysis and an unmatched multiple-tests analysis.

**Setting:** Nationally centralized database of Maccabi Healthcare Services (MHS), an Israeli national health fund that covers 2.5 million people.

**Participants:** The study population included 97,499 MHS members aged 60 or older who were eligible to receive a fourth vaccine dose and performed at least one PCR test during the study period. Of them, 27,876 received the fourth dose and 69,623 received only three doses.

**Main outcomes and measures:** Analyses focused on the period from January 10, 2022 (7 days after the fourth dose was first administered to eligible individuals) to March 13, 2022, an Omicron-dominant period in Israel. We evaluated two SARS-CoV-2-related outcomes: (1) breakthrough infection, defined as a positive PCR test performed 7 or more days after inoculation with the BNT162b2 vaccine; and (2) breakthrough infection resulting in a severe disease, defined as COVID-19-related hospitalization or COVID-19 associated mortality.

**Results:** A fourth dose provided considerable additional protection against both SARS-CoV-2 infection and severe disease relative to three doses of the vaccine. However, vaccine effectiveness against infection varied over time, peaking during the third week with a VE of 64% (95% CI: 62.0%-65.9%) and declining to 29.2% (95% CI: 17.7%-39.1%) by the end of the 10-week follow-up period. Unlike VE against infection, the relative effectiveness of a fourth dose against severe COVID-19 was maintained at high level (>73%) throughout the 9-week follow-up period. Importantly, severe disease was a relatively rare event, occurring in <1% of both fourth dose and third dose only recipients.

**Conclusions:** A fourth dose of the BNT162b2 vaccine provided considerable additional protection against both SARS-CoV-2 infection and severe disease relative to three doses of the vaccine. However, effectiveness of the fourth dose against infection wanes sooner than that of the third dose.

## Introduction

The surge of infections with the Delta (B.1.617.2) variant of SARS-CoV-2 during the summer of 2021,^1^ combined with a growing body of evidence demonstrating waning of BioNTech/Pfizer mRNA BNT162b2 vaccine-induced immunity^2–5^ led Israel to launch a national third-dose (booster) vaccination campaign in August 2021. The booster was shown to largely restore short-term effectiveness,^6–8^ and by December 31, 2021, over 4.2 million individuals (∼45% of individuals of all ages) received a booster shot.^1^ However, the rapid spread of the Omicron variant (B.1.1.529) from late December through March 2022^9^ caused a sharp increase in infection rates and COVID-19-related hospitalizations.^1^ The rising incidence alongside evidence of a relatively rapid decrease in protection conferred by the third dose^10,11^ prompted Israel’s decision to administer a fourth dose (a second booster). Recommendations were first given for immunosuppressed individuals on December 30, 2021,^12^ and 3 days later for all individuals aged 60 or older and those at high risk of exposure (e.g. healthcare workers). Eligible persons must have received the third dose at least four months prior to the second booster shot.

Thus far, sufficient real-world evidence demonstrating the effectiveness of a fourth dose against infection and severe COVID-19 are lacking. Leveraging the centralized database of Maccabi Healthcare Services (MHS), an Israeli national health fund that covers 2.5 million people, we examined the short-term effectiveness of a fourth dose over the span of 10 weeks.

## Methods

### Data sources and data extraction

MHS, which covers 26.7% of the population and provides a representative sample of the Israeli population, has maintained a centralized database of electronic medical records (EMRs) for three decades, with less than 1% disengagement rate among its members, allowing for comprehensive longitudinal medical follow-up, and includes demographic data, measurements, inpatient and outpatient procedures and diagnoses, medications, imaging records and laboratory data from a single central laboratory.

Anonymized EMRs were retrieved from MHS’s centralized computerized database. Individual-level data for the study population included age, biological sex, socioeconomic status (SES) index, a coded geographical statistical area (GSA; the smallest geostatistical unit assigned by Israel’s National Bureau of Statistics, which roughly correspond to neighborhoods), and whether a person currently resides at a nursing home or an assisted living residence. The SES index was measured on a scale from 1 (lowest) to 10 based on several parameters including household income, educational qualifications, household crowding, and car ownership. Data collected also encompassed the last documented body mass index (BMI) (where obesity was defined as BMI ≥ 30) and information on chronic diseases from MHS’s automated registries, including cardiovascular diseases,^13^ hypertension,^14^ diabetes,^15^ chronic kidney disease (CKD),^16^ chronic obstructive pulmonary disease (COPD), and immunocompromised conditions. SARS-CoV-2-related information included dates of vaccinations and results of any PCR tests for SARS-CoV-2 (including tests performed within and outside of MHS) and COVID-19-related hospitalization dates.

### Study population

The study population included all MHS members aged 60 or older who were eligible to receive a fourth vaccine dose on January 3, 2022, the first day of eligibility according to the Israeli Ministry of Health. Eligible individuals were those who received at least three doses of the vaccine and for whom, on January 1, at least four months had passed since the third (booster) dose. We excluded individuals who had a positive SARS-CoV-2 polymerase chain reaction (PCR) test prior to the start of the study period (January 10, 2022) and persons with a possibly incomplete COVID-19-related medical history during the pandemic, i.e. those who joined MHS after March 2020.

### Design and statistical analysis

Analyses focused on the period from January 10, 2022 (7 days after the fourth dose was first administered to eligible individuals) to March 13, 2022, an Omicron-dominant period in Israel.^9^ We evaluated two SARS-CoV-2-related outcomes: (1) breakthrough infection, defined as a positive PCR test performed 7 or more days after inoculation with the BNT162b2 vaccine, where the 7-day cutoff was based on previous breakthrough infection definitions in vaccine effectiveness studies;^6,17^ and (2) breakthrough infection resulting in a severe disease, defined as COVID-19-related hospitalization (up to one month from the first positive PCR test) or COVID-19 associated mortality.

Prior SARS-CoV-2 studies demonstrated a time-dependent increase (and then decrease) in the level of protection conferred by the vaccine. Therefore, we stratified the analysis by time-since-vaccination in equal-day intervals (7-13, 14-20, 21-27, 28-34, 35-41, 42-48, 49-55, 56-62, 63-69 days), where the aim was to estimate the reduction in the odds of a positive outcome at these different time points after inoculation with a fourth dose. The vaccination status was determined at the time of the PCR test or the COVID-19 hospitalization or mortality, the earliest of the three. For the outcome of severe disease, we stratified by three-weeks intervals of time-since-vaccination, due to insufficient numbers of severe outcomes.

For our primary analysis, we used a test-negative case-control design, which has been strongly argued for in COVID-19 studies due to its ability to better control for bias stemming from healthcare seeking and testing behavior.^6,18,19^ We defined cases as (1) individuals with a SARS-CoV-2-positive PCR test during the study period, or (2) individuals diagnosed with severe COVID-19 during the study period; cases were defined separately for each outcome. Eligible controls were individuals with a negative PCR test result who had not tested positive previously. Each outcome was evaluated with two alternative analytic approaches: a matched analysis and an unmatched, multiple-tests analysis.

### Matched analysis

The matched approach consisted of a 1:1 matching based on sex, GSA, calendrical week of testing (to account for potential time-varying risk within the outcome period), the month of receipt of the third dose (to mitigate possible bias relating to waning of the third dose), and a categorical variable relating to the living environment (a medical nursing home, an assisted living facility, or a private residence). The latter was included in light of early-pandemic surges in these environments, which led to differential regulations in these institutions, mandating staff and residents to get vaccinated and limiting visits from non-residents. Therefore, exposure was substantially different in these facilities compared to the rest of the population.

The first positive PCR test (or first COVID-19-associated hospitalization or mortality in the severe disease analysis) and the first negative PCR test were the only tests included for each case and control, respectively.^20^ Negative tests for cases were excluded, rendering participants as either cases or controls, but not both. The rationale was to avoid a potential bias stemming from repeated tests, indicating different healthcare-seeking behavior and potentially a lower pre-test risk of infection.^20^

A conditional logistic regression model that accounted for the matching was fit to the data. The marginal vaccine effectiveness (VE)^6^ of the fourth dose (compared to the third dose) was calculated as 100%*[1-(Odds Ratio)] for each of week-since-vaccination interval. The odds ratio estimated the relative, or *multiplicative*, effect of the fourth dose compared to the third dose, rather than the absolute effectiveness compared to being unvaccinated; the latter analysis is not possible given the rapid rollout and high vaccination coverage among older individuals in Israel.

To address potential confounders, we adjusted for underlying comorbidities, including obesity, cardiovascular diseases, diabetes, hypertension, chronic kidney disease, COPD and immunosuppression conditions. Additionally, we adjusted for age, using a binary cutoff of 70 years or older, as previous studies have indicated older age groups are at higher risk, especially for severe disease. We also adjusted for a categorical variable consisting of the number of PCR tests each person undertook from the beginning of the pandemic (March 2020) until the start of the outcome period, as a proxy for SARS-CoV-2-related healthcare-seeking behavior. The covariate included the entire pandemic period apart from the outcome period, in order to differentiate this *behavioral* variable from the dependent variable.^5,21^

### Multiple tests unmatched analysis

In the unmatched approach to the test-negative design, we allowed persons to contribute multiple negative tests, but excluded them once they tested positive or became hospitalized with COVID-19.^6,11^ We adjusted for all of the covariates included in the matched approach (i.e., comorbidities, age group, and number of previous PCR tests) as well as some of the previously matched-upon parameters, namely biological sex, calendrical week of testing, residential socioeconomic status, the month of receipt of the third dose and residence in a nursing home or assisted living facility. In this unmatched approach, a Generalized Estimating Equation (GEE) logistic regression model was fit to the data in order to account for the possibility of repeated samples from the same participant,^22^ assuming an exchangeable correlation structure. The marginal vaccine effectiveness was estimated in the same way as the matched analysis.

All analyses were performed using R Studio version 3.6 with the MatchIt, gee, geepack and survival packages.

## Data Availability

According to the Israel Ministry of Health regulations, individual-level data cannot be shared openly. Specific requests for remote access to de-identified community-level data should be referred to KSM, Maccabi Healthcare Services Research and Innovation Center.

## Ethics declaration

This study was approved by the MHS (Maccabi Healthcare Services) Institutional Review Board. Due to the retrospective design of the study, informed consent was waived by the IRB, as all identifying details of the participants were removed before computational analysis.

## Code availability statement

Specific requests for remote access to the code used for data analysis should be referred to KSM, Maccabi Healthcare Services Research and Innovation Center.

## Results

There were 97,499 MHS members over the age of 60 who were eligible for the study and performed at least one PCR test during the outcome period of January 10 to March 13, 2022. Within that time period, 233,582 PCR tests were performed by 27,876 participants who received a fourth dose of BNT162b2 and 69,623 persons who were eligible for a fourth dose but had not (yet) received one. Population characteristics are described in **Table 1**. The recipients of the fourth dose were overall more chronically ill, possibly correlated to targeted vaccination campaigns and stronger compliance to recommendations. These differences stress the need for the performed adjustment.

**Table 1.** Demographic characteristics of individuals with at least 3 doses of vaccine who were tested between January 10, 2022 – March 13, 2022

|  | 3-dose vaccinated<br>(*N*=69623) | 4-dose vaccinated<br>(*N*=27876) | Overall<br>(*N*=97499) |
| --- | --- | --- | --- |
| **Sex** |  |  |  |
| Female | 37777 (54.3%) | 15583 (55.9%) | 53360 (54.7%) |
| Male | 31846 (45.7%) | 12293 (44.1%) | 44139 (45.3%) |
| **Age** |  |  |  |
| Mean (SD) | 70.1 (7.63) | 72.6 (8.66) | 70.8 (8.02) |
| Median [Min, Max] | 68.8 [60.0, 104] | 71.2 [60.0, 108] | 69.4 [60.0, 108] |
| **SES** |  |  |  |
| High (7-10) | 29759 (42.7%) | 10723 (38.5%) | 40482 (41.5%) |
| Middle (4-6) | 31814 (45.7%) | 13243 (47.5%) | 45057 (46.2%) |
| Low (1-3) | 8050 (11.6%) | 3910 (14.0%) | 11960 (12.3%) |
| **CVD** |  |  |  |
| No | 51897 (74.5%) | 19807 (71.1%) | 71704 (73.5%) |
| Yes | 17726 (25.5%) | 8069 (28.9%) | 25795 (26.5%) |
| **DM** |  |  |  |
| No | 53004 (76.1%) | 20151 (72.3%) | 73155 (75.0%) |
| Yes | 16619 (23.9%) | 7725 (27.7%) | 24344 (25.0%) |
| **Hypertension** |  |  |  |
| No | 31704 (45.5%) | 11034 (39.6%) | 42738 (43.8%) |
| Yes | 37919 (54.5%) | 16842 (60.4%) | 54761 (56.2%) |
| **CKD** |  |  |  |
| No | 46043 (66.1%) | 16366 (58.7%) | 62409 (64.0%) |
| Yes | 23580 (33.9%) | 11510 (41.3%) | 35090 (36.0%) |
| **COPD** |  |  |  |
| No | 65301 (93.8%) | 25929 (93.0%) | 91230 (93.6%) |
| Yes | 4322 (6.2%) | 1947 (7.0%) | 6269 (6.4%) |
| **Obesity (BMI ≥30)** |  |  |  |
| No | 53598 (77.0%) | 20904 (75.0%) | 74502 (76.4%) |
| Yes | 16025 (23.0%) | 6972 (25.0%) | 22997 (23.6%) |
| **Immunosuppression** |  |  |  |
| No | 60690 (87.2%) | 23740 (85.2%) | 84430 (86.6%) |
| Yes | 8933 (12.8%) | 4136 (14.8%) | 13069 (13.4%) |
| **Assisted Living** |  |  |  |
| No | 68892 (99.0%) | 27318 (98.0%) | 96210 (98.7%) |
| Yes | 731 (1.0%) | 558 (2.0%) | 1289 (1.3%) |
| **Nursing Home** |  |  |  |
| No | 68078 (97.8%) | 26182 (93.9%) | 94260 (96.7%) |
| Yes | 1545 (2.2%) | 1694 (6.1%) | 3239 (3.3%) |
| **Test-taking behaviors** |  |  |  |
| 0 prev. tests | 12507 (18.0%) | 4502 (16.2%) | 17009 (17.4%) |
| 1-2 prev. tests | 21295 (30.6%) | 7569 (27.2%) | 28864 (29.6%) |
| 3>= prev. tests | 35821 (51.4%) | 15805 (56.7%) | 51626 (53.0%) |
| **Third dose month** |  |  |  |
| 2021-8 | 64172 (92.2%) | 26924 (96.6%) | 91096 (93.4%) |
| 2021-9 | 4407 (6.3%) | 849 (3.0%) | 5256 (5.4%) |
| 2021-10 | 995 (1.4%) | 103 (0.4%) | 1098 (1.1%) |
| 2021-11 | 49 (0.1%) | 0 (0%) | 49 (0.1%) |

SD – Standard Deviation; SES – Socioeconomic status on a scale from 1 (lowest) to 10; DM – Diabetes Mellitus; CKD – Chronic Kidney Disease; COPD – Chronic Obstructive Pulmonary Disease; BMI – Body Mass Index.

### Effectiveness against breakthrough infections

The unmatched multiple-test analysis included 229,433 PCR tests, of which 35,101 (15.3%) were positive. The matched test-negative analysis included 25,536 cases and their matched controls (**Table S1**). Marginal vaccine effectiveness estimates by week since receipt of the fourth dose are detailed in **Table 2**. The adjusted odds ratios (aOR) from which effectiveness estimates were derived can be found in **Tables S3-S4**.

**Table 2.** Adjusted fourth dose marginal vaccine effectiveness (VE) against SARS-CoV-2 infection. VE was defined as 100%*[1-(Odds Ratio)] for each of the week-since-vaccination intervals. The reference group is those who only received three doses of BNT162b2. ORs were adjusted for comorbidities, age group, nursing home or assisted living residence, previous test-taking behavior, biological sex, calendrical week of testing, residential socioeconomic status, the month of receipt of the third dose.

| **Time after fourth**<br>**dose receipt** | **Multiple tests analysis** | **Matched analysis** |
| --- | --- | --- |
| 7-13 d | 45.97% (43.66%, 48.19%) | 56.05% (53.41%, 58.53%) |
| 14-20 d | 61.79% (59.89%, 63.6%) | 64.29% (61.73%, 66.68%) |
| 21-27 d | 64.00% (62.04%, 65.85%) | 63.02% (60.03%, 65.78%) |
| 28-34 d | 61.02% (58.62%, 63.28%) | 59.51% (55.63%, 63.05%) |
| 35-41 d | 58.80% (55.73%, 61.65%) | 56.32% (50.94%, 61.11%) |
| 42-48 d | 57.11% (53.36%, 60.56%) | 49.02% (41.04%, 55.92%) |
| 49-55 d | 52.77% (48.17%, 56.97%) | 41.93% (31.56%, 50.72%) |
| 56-62 d | 42.50% (36.41%, 48%) | 41.50% (29.58%, 51.41%) |
| 63-69 d | 29.20% (17.74%, 39.07%) | 27.08% (4.24%, 44.47%) |

The fourth-dose marginal effectiveness against infection largely increased in the second week after inoculation until peaking during the third week, with a VE of 64% (95% confidence interval [CI]: 62.04%, 65.85%) compared to those vaccinated with only three doses in the multiple-tests analysis and similar results for the matched analysis (**Table 2**). However, vaccine effectiveness began to decline four weeks after inoculation, with relative effectiveness on week 8 roughly dropping back to levels observed during the first week, and reaching 29.2% (95% CI: 17.74%, 39.07%) by week 9. Similar results were obtained with the matched analysis, although fewer observations led to wide confidence intervals during the last week.

Comorbidities were not significantly associated with the odds of infection (**Table S3-S4**). However, the odds of infection were lower among those 70 years or older in both models. Likewise, having received three or more PCR tests prior to the study period was associated with a significantly reduced odds of infection. The unmatched multiple-test analysis highlighted the importance of controlling for test week, where the odds of infection were highest in late January-early February. Additionally, residing in a nursing home or an assisted living facility was associated with a significant protective effect, with an OR of 0.55 (95% CI: 0.51, 0.59) and 0.62 (95% CI: 0.54, 0.71), respectively.

### Effectiveness against severe disease

There were 194,906 cases and controls included in the multiple-test analysis of severe disease, of which 574 (0.3%) were either hospitalized or died as a result of COVID-19; 406 cases and their matched controls were included in the matched analysis (**Table S2**). There were 106 deaths during the follow-up period, of which 77 were among third-dose-only recipients, and 23 were among fourth-dose vaccinees during the first three weeks after inoculation.

Unlike VE against infection, the VE against severe disease did not vary significantly as a function of time-since-vaccination (**Table 3, Figure 2)**. The marginal effectiveness of the fourth dose compared to a third dose was 86.1% (95% CI: 73.3%, 92.8%) during weeks 6-9 in the multiple-tests analysis.

**Table 3.** Adjusted fourth dose marginal vaccine effectiveness (VE) against severe COVID-19. VE was defined as 100%*[1-(Odds Ratio)] for each time-since-vaccination interval. The reference group is those who only received three doses of BNT162b2. ORs were adjusted for comorbidities, age group, nursing home or assisted living residence, previous test-taking behavior, biological sex, calendrical week of testing, residential socioeconomic status, the month of receipt of the third dose.

| **Time after fourth dose receipt** | **Multiple tests analysis** | **Matched analysis** |
| --- | --- | --- |
| **7-27 d** | 73.4% (66.46%, 78.97%) | 82.5% (70.48%, 89.59%) |
| **28-48 d** | 73.8% (64.26%, 80.82%) | 70.3% (37.37%, 85.9%) |
| **49-69 d** | 86.1% (73.3%, 92.77%) | 87.1% (0%, 98.36%) |

**Figure 1.**
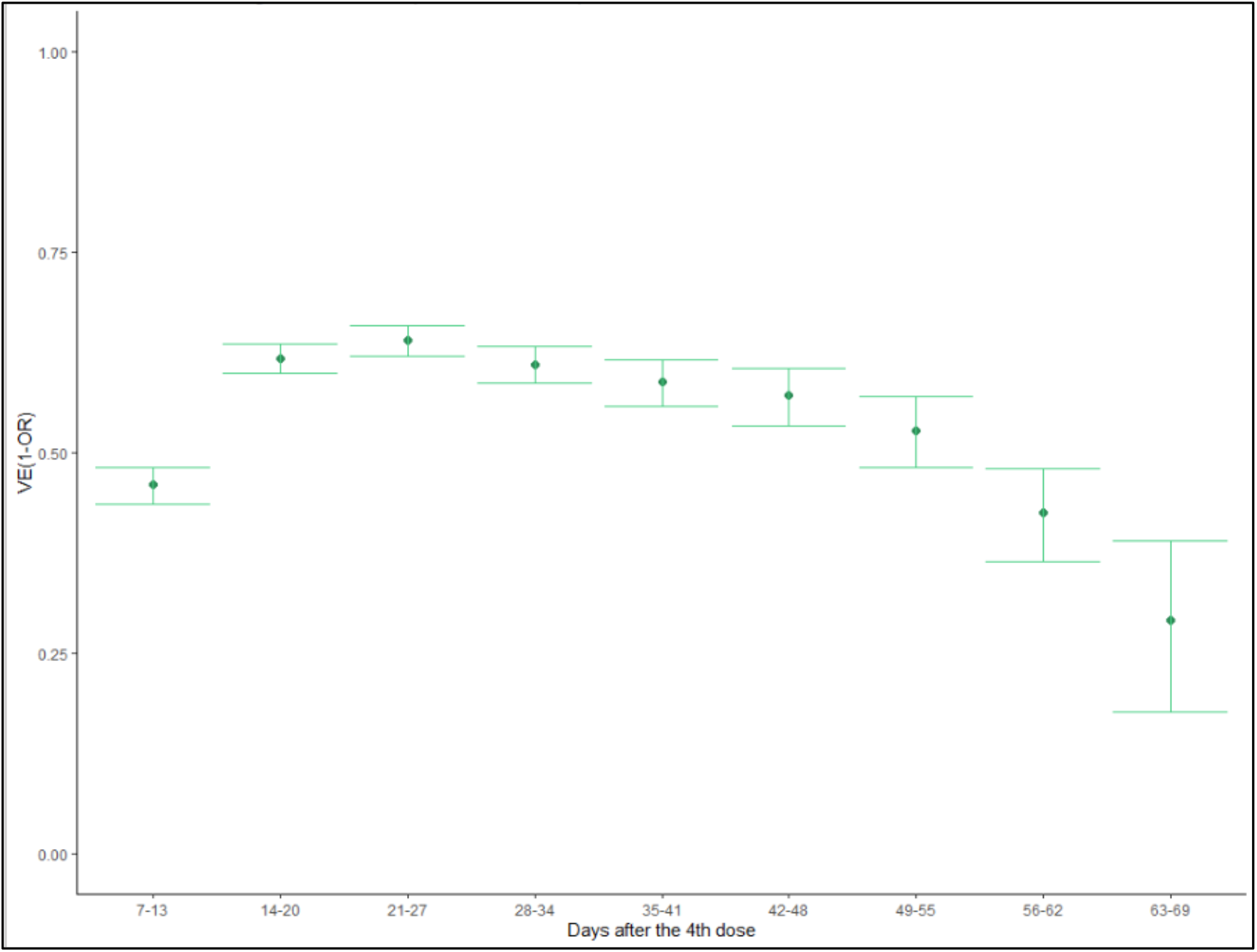
Adjusted fourth dose vaccine effectiveness against SARS-CoV-2 infection relative to three doses. Multiple tests approach.

**Figure 2.**
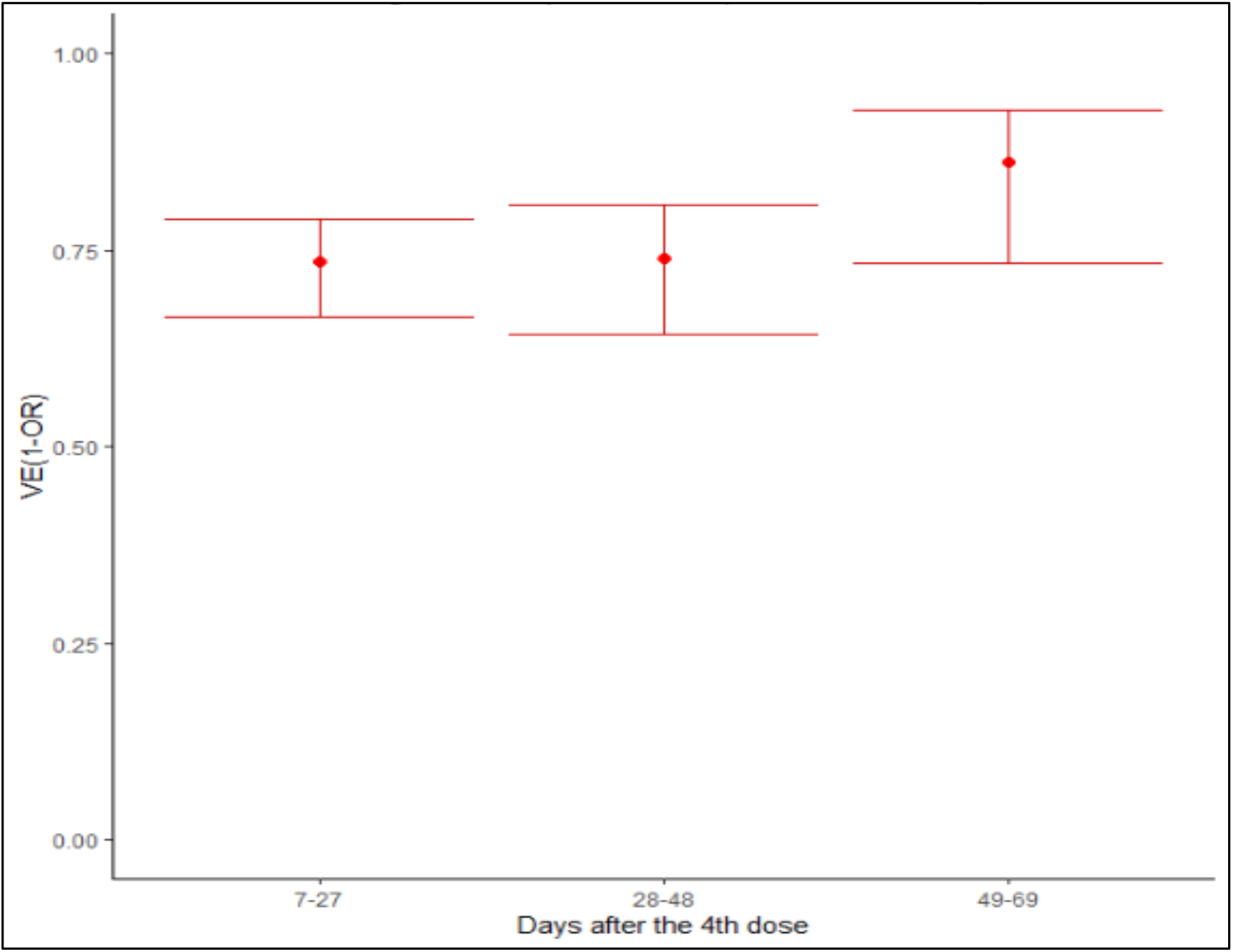
Adjusted fourth dose vaccine effectiveness against SARS-CoV-2 severe disease relative to three doses. Multiple tests approach.

Individuals aged 70 years or older had a significantly higher odds of severe disease, with an OR of 2.34 (95% CI: 1.87, 2.92). Comorbidities were also associated with an increased odds of severe diseases as well (**Tables S5-S6**). The relative paucity of observations limited the power in the matched analysis (**Table 3, Tables S5-S6, Figure S2**).

## Discussion

This study investigated the relative effectiveness of a fourth dose of the BioNTech/Pfizer mRNA BNT162b2 vaccine compared to the third dose, against both infection with the SARS-CoV-2 Omicron variant and infection resulting in a severe COVID-19 disease, assessed by hospitalizations and mortality, among individuals 60 years of age and older in Israel. Using a test-negative design and performing both a matched analysis and an unmatched multiple-tests analysis, we found a fourth dose provided considerable additional protection against both SARS-CoV-2 infection and severe disease relative to three doses of the vaccine. However, vaccine effectiveness against infection varied over time, peaking during the third week with a VE of 64% (95% CI: 62.0%-65.9%) and declining to 29.2% (95% CI: 17.7%-39.1%) by the end of the 10-week follow-up period.

The waning of vaccine effectiveness against infection is consistent with previous observations for the second and third doses of the BNT162b2b vaccine.^10,11^ Nonetheless, compared to the previously demonstrated waning pattern of the relative effectiveness of three doses compared to two doses in real-world settings – which begins around 3 months after inoculation – it appears that effectiveness of the fourth dose wanes sooner, similarly to the fact that the third dose wants sooner than the second dose.^23,11^ This more rapid decline is potentially explained by a reduced effectiveness of the BNT162b2b vaccine against the Omicron variant.^24,25^ However, comparison of variants in relation to waning protection of sequential vaccine doses cannot be studied today in most Western countries, due to real-world circulation of variants. Additionally, we need to consider that frequent stimulation with the same mRNA vaccine triggers an immunological response that is yet to be fully understood in terms of duration, effectiveness and patterns of waning when exposed to different variants.

Unlike VE against infection, the relative effectiveness of a fourth dose against severe COVID-19 was maintained at high level (>73%) throughout the 9-week follow-up period, a sustained effect against severe disease that demonstrated in previous doses as well.^20,26^ Importantly, it should be noted that severe disease was a relatively rare event, occurring in <1% of both fourth dose and third dose only recipients.

Our analysis is subject to a number of limitations. First, in order to provide timely evidence of the effectiveness of a fourth dose of BNT162b2b vaccine, we were only able to include 10 weeks of data. Although the pattern of a short-term increase in protection against infection followed by waning is already present, long-term effectiveness needs to be evaluated. This is particularly important for estimates of effectiveness of the fourth dose against severe disease. It has been previously demonstrated that protection from previous doses against severe disease wanes more slowly compared to waning of protection against infection.^20,26^ Nonetheless, our study suggests a more rapid waning of protection against infection from a fourth dose compared to previous doses; therefore, waning of effectiveness against COVID-19-associated hospitalization and mortality needs to be further examined over a longer period.

A second inherent limitation stems from the varying dominance of different SARS-CoV-2 variants over time. The post-fourth dose period in Israel has been dominated by the Omicron variant, which renders it difficult to assess the relative effectiveness of the fourth dose against other variants, a well-recognized limitation of real-world analyses during this pandemic.^6,27,28^ Furthermore, as the eligible population for a fourth dose was comprised of individuals aged 60 or older, we cannot infer effectiveness and potential waning in younger persons. Additionally, the fourth-dose recipients were overall sicker, a fact that matches targeted vaccination campaigns and previous rollout policies. Adjusting for comorbidities by virtue of an available comprehensive medical history, as well as adjustment and matching by other factors including timing of the third vaccine dose, residential and social factors, and previous testing renders residual confounding less likely.

Lastly, an important limitation relates to the observational nature and the real-world data collection of this study, in particular the lack of pre-defined PCR testing protocols implemented in the study population. This limitation has been elaborately discussed in previous COVID-19 observational studies, and could lead to potential biases relating to healthcare-seeking behavior.^6,11,29,30^ The test-negative design, which does not assume those who are not tested are eligible to serve as uninfected controls, aims to somewhat mitigate this potential bias. Furthermore, some studies have included rapid antigen tests in their analysis, treating them equally to PCR tests, whereas this study did not. While we admittedly have fewer observations by excluding rapid antigen tests, such tests are generally considered less reliable, and negative at-home tests are not reported. Furthermore, a PCR-test-for-all policy was in place for the examined age group (60 years or older, eligible for a fourth dose) during the follow-up period, making testing accessible to the study population.

## Conclusions

In conclusion, this study demonstrated a significant additional protection against both SARS-CoV-2 infection and severe disease relative to three doses of the vaccine. However, vaccine effectiveness against infection varied over time, peaking during the third week with a VE of 64% and declining to 29.2% by the end of the 10-week follow-up period. Thus, effectiveness of the fourth dose against infection wanes sooner than that of the third dose. Unlike VE against infection, the relative effectiveness of a fourth dose against severe COVID-19 was maintained at high level (>73%) throughout the 9-week follow-up period, though severe disease was a relatively rare event, occurring in <1% of both fourth dose and third dose only recipients.

## Supplementary Tables and Figures

**Table S1.** Testing results among those with at least three doses of the vaccine at different time points, January 10, 2021-March 13, 2022

|  | Multiple tests<br>approach |  | 1:1 matched<br>approach |  |
| --- | --- | --- | --- | --- |
|  | No. |  |  |  |
| Time after fourth<br>dose receipt | Positive | Negative | Positive | Negative |
| Received only<br>three doses | 22046 | 74268 | 15730 | 10839 |
| 7-13 d | 3555 | 23423 | 2941 | 4576 |
| 14-20 d | 2439 | 19478 | 1849 | 3152 |
| 21-27 d | 2033 | 17484 | 1570 | 2467 |
| 28-34 d | 1613 | 15274 | 1208 | 1731 |
| 35-41 d | 1084 | 12664 | 750 | 1027 |
| 42-48 d | 796 | 11146 | 524 | 644 |
| 49-55 d | 654 | 9719 | 422 | 488 |
| 56-62 d | 625 | 8214 | 373 | 448 |
| 63-69 d | 256 | 2662 | 169 | 164 |
| Overall | 35101 | 194332 | 25536 | 25536 |

**Table S2.**
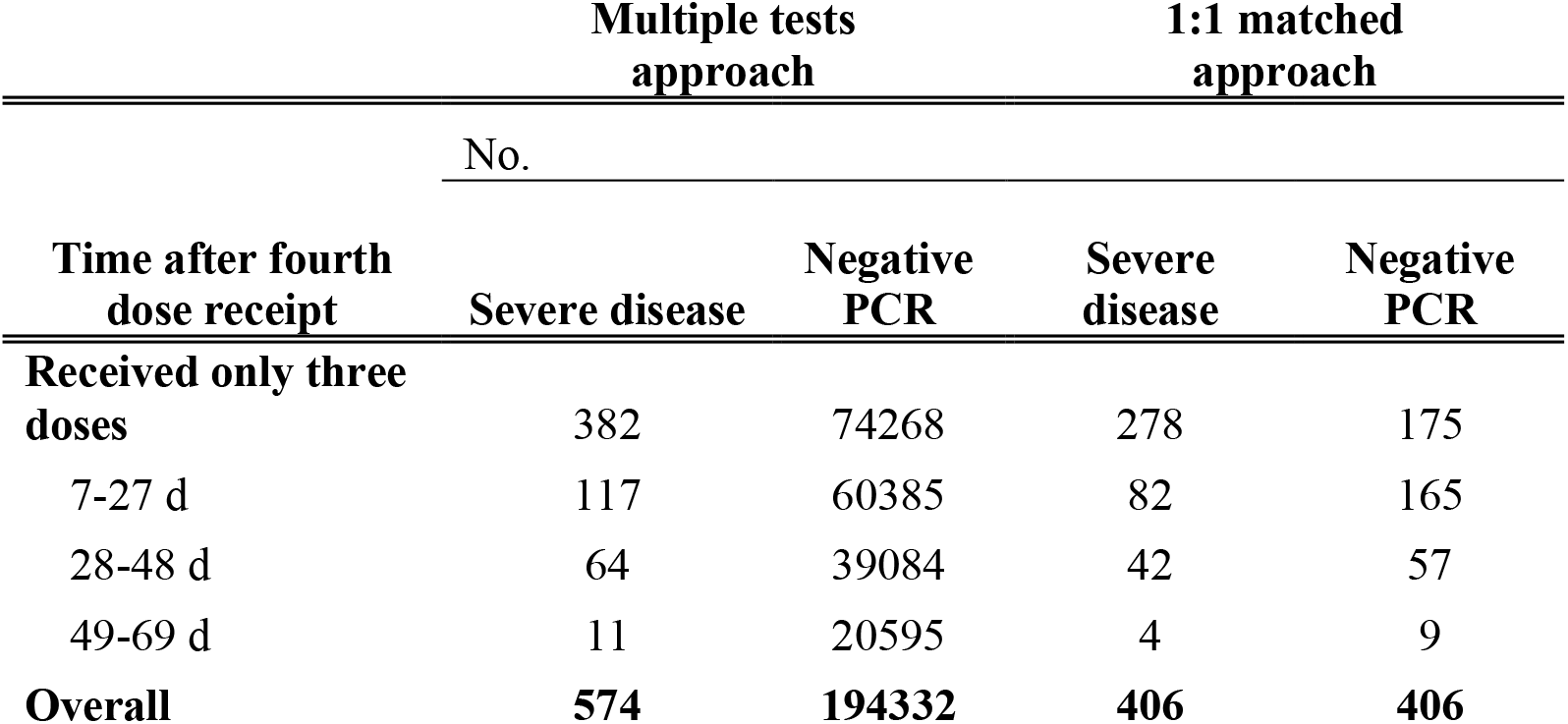
Tests and severe disease outcomes among those with at least three doses of the vaccine at different time points, January 10, 2022-March 13, 2022

**Table S3.** Odds ratios (OR) for SARS-CoV-2 infection, multiple tests analysis.

| Variable | Category | OR | 95% CI |
| --- | --- | --- | --- |
| **Vaccination status** |  |  |  |
|  | Three dose only vaccinees | Ref |  |
|  | Four dose vaccinees, days 7-13 | 0.540 | (0.36, 0.4) |
|  | Four dose vaccinees, days 14-20 | 0.382 | (0.34, 0.38) |
|  | Four dose vaccinees, days 21-27 | 0.360 | (0.37, 0.41) |
|  | Four dose vaccinees, days 28-34 | 0.390 | (0.38, 0.44) |
|  | Four dose vaccinees, days 35-41 | 0.412 | (0.39, 0.47) |
|  | Four dose vaccinees, days 42-48 | 0.429 | (0.43, 0.52) |
|  | Four dose vaccinees, days 49-55 | 0.472 | (0.52, 0.64) |
|  | Four dose vaccinees, days 56-62 | 0.575 | (0.61, 0.82) |
|  | Four dose vaccinees, days 63-69 | 0.708 | (0.52, 0.56) |
| **Sex** |  |  |  |
|  | Female | Ref |  |
|  | Male | 1.26 | (1.23, 1.3) |
| **Age group, yr.** |  |  |  |
|  | 60-69 | Ref |  |
|  | ≥70 | 0.81 | (0.79, 0.83) |
| **SES** |  |  |  |
|  | Low (1-3) | 0.99 | (0.95, 1.02) |
|  | Medium (4-6) | Ref |  |
|  | High (7-10) | 0.89 | (0.87, 0.92) |
| **Comorbidities** |  |  |  |
|  | Cardiovascular disease | 1.00 | (0.98, 1.03) |
|  | Diabetes mellitus | 1.00 | (0.97, 1.03) |
|  | Hypertension | 1.01 | (0.98, 1.04) |
|  | Chronic Kidney Disease | 0.95 | (0.92, 0.97) |
|  | Chronic Obstructive Pulmonary Disease | 0.93 | (0.89, 0.98) |
|  | Immunosuppression | 1.00 | (0.97, 1.04) |
|  | Obesity (BMI≥30) | 1.01 | (0.98, 1.04) |
| **Living environment** |  |  |  |
|  | Independent housing | Ref |  |
|  | Assisted living | 0.62 | (0.54, 0.71) |
|  | Nursing home | 0.55 | (0.51, 0.59) |
| **Test Week** |  |  |  |
|  | January 10-16 (week 1) | Ref |  |
|  | January 17-23 (week 2) | 1.45 | (1.39, 1.51) |
|  | January 23-30 (week 3) | 1.64 | (1.57, 1.71) |
|  | January 31-February 6 (week 4) | 1.49 | (1.42, 1.56) |
|  | February 7-13 (week 5) | 1.21 | (1.15, 1.27) |
|  | February 14-20 (week 6) | 0.90 | (0.85, 0.96) |
|  | February 21-27 (week 7) | 0.76 | (0.71, 0.81) |
|  | February 28-March 6 (week 8) | 0.67 | (0.62, 0.72) |
|  | March 7-March 13 (week 9) | 0.63 | (0.58, 0.69) |
| **Month of receipt of third dose** |  |  |  |
|  | August 2021 | Ref |  |
|  | September-November 2021 | 0.92 | (0.88, 0.97) |
| **Previous tests** |  |  |  |
|  | 0 | Ref |  |
|  | 1-2 | 0.79 | (0.76, 0.81) |
| | $\geq 3$ | 0.42 | (0.41, 0.43) |

OR – Odds Ratio.

**Table S4.** Odds ratios (OR) for SARS-CoV-2 infection, matched analysis.

| Variable | Category | OR | 95% CI |
| --- | --- | --- | --- |
| **Vaccination status** |  |  |  |
|  | Three dose only vaccinees | Ref |  |
|  | Four dose vaccinees, days 7-13 | 0.440 | (0.41, 0.47) |
|  | Four dose vaccinees, days 14-20 | 0.357 | (0.33, 0.38) |
|  | Four dose vaccinees, days 21-27 | 0.370 | (0.34, 0.4) |
|  | Four dose vaccinees, days 28-34 | 0.405 | (0.37, 0.44) |
|  | Four dose vaccinees, days 35-41 | 0.437 | (0.39, 0.49) |
|  | Four dose vaccinees, days 42-48 | 0.510 | (0.44, 0.59) |
|  | Four dose vaccinees, days 49-55 | 0.581 | (0.49, 0.68) |
|  | Four dose vaccinees, days 56-62 | 0.585 | (0.49, 0.7) |
|  | Four dose vaccinees, days 63-69 | 0.729 | (0.56, 0.96) |
| **Age group, yr.** |  |  |  |
|  | 60-69 | Ref |  |
|  | ≥70 | 0.829 | (0.8, 0.86) |
| **Comorbidities** |  |  |  |
|  | Cardiovascular disease | 1.019 | (0.97, 1.07) |
|  | Diabetes mellitus | 1.006 | (0.96, 1.05) |
|  | Hypertension | 0.990 | (0.95, 1.03) |
|  | Chronic Kidney Disease | 0.987 | (0.95, 1.03) |
|  | Chronic Obstructive Pulmonary Disease | 0.900 | (0.83, 0.97) |
|  | Immunosuppression | 0.993 | (0.94, 1.05) |
|  | Obesity (BMI≥30) | 0.977 | (0.94, 1.02) |
| **Previous tests** |  |  |  |
|  | 0 | Ref |  |
|  | 1-2 | 0.917 | (0.87, 0.96) |
|  | ≥3 | 0.777 | (0.74, 0.82) |

OR – Odds Ratio.

**Table S5.**
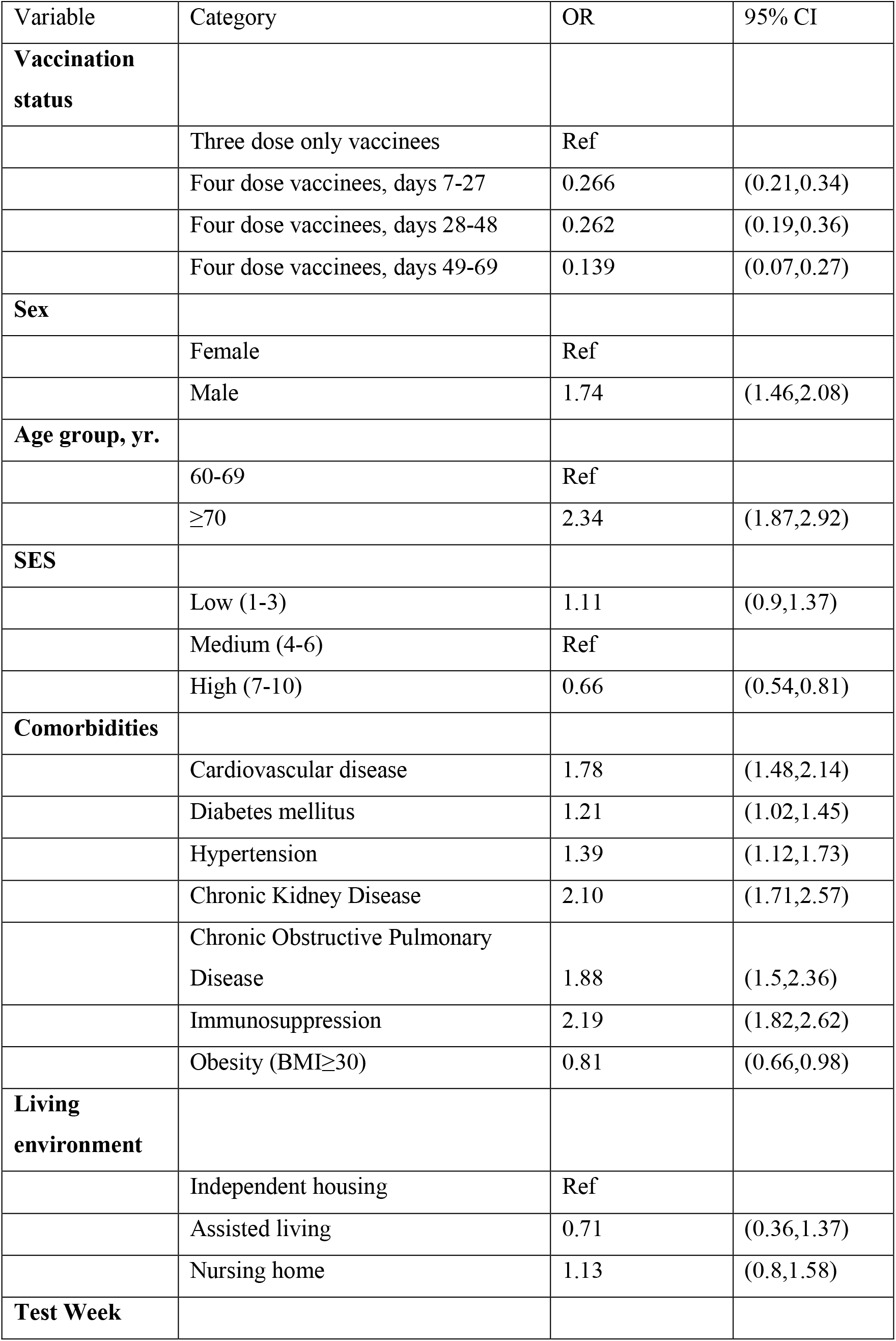
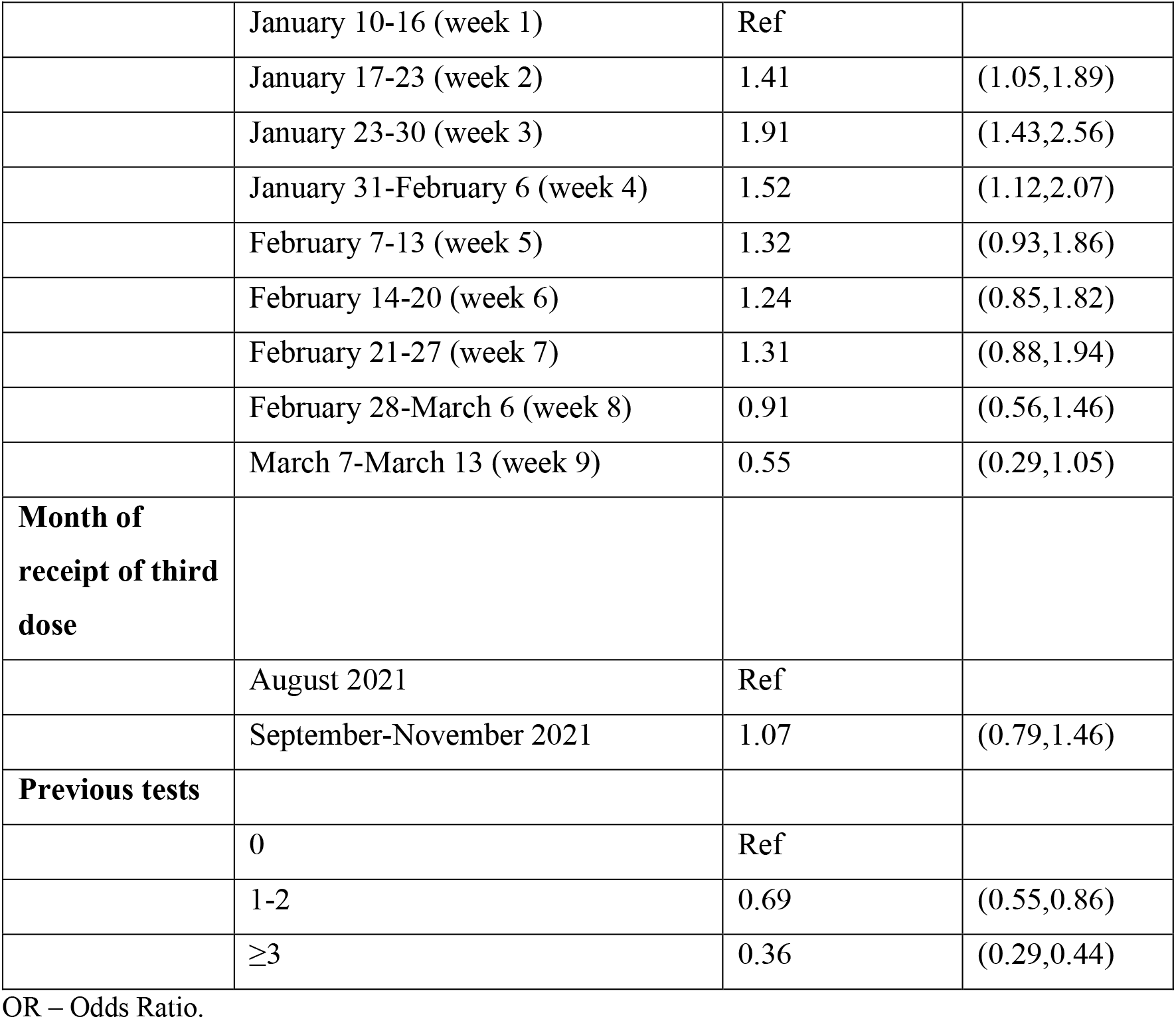
Odds ratios (OR) for SARS-CoV-2 associated severe disease, multiple tests analysis.

**Table S6.** Odds ratios (OR) for SARS-CoV-2 associated severe disease, matched tests analysis.

| Variable | Category | OR | 95% CI |
| --- | --- | --- | --- |
| **Vaccination status** |  |  |  |
|  | Three dose only vaccinees | Ref |  |
|  | Four dose vaccinees, days 7-27 | 0.175 | (0.1, 0.3) |
|  | Four dose vaccinees, days 28-48 | 0.297 | (0.14, 0.63) |
|  | Four dose vaccinees, days 49-69 | 0.129 | (0.02, 1.01) |
| **Age group, yr.** |  |  |  |
|  | 60-69 | Ref |  |
| | $\geq 70$ | 2.324 | (1.52, 3.56) |
| **Comorbidities** |  |  |  |
|  | Cardiovascular disease | 2.064 | (1.33, 3.2) |
|  | Diabetes mellitus | 1.169 | (0.77, 1.77) |
|  | Hypertension | 1.603 | (1.01, 2.53) |
|  | Chronic Kidney Disease | 2.605 | (1.67, 4.06) |
|  | Chronic Obstructive Pulmonary Disease | 1.663 | (0.84, 3.28) |
|  | Immunosuppression | 2.019 | (1.28, 3.18) |
| | Obesity (BMI$\geq 30$) | 0.584 | (0.37, 0.93) |
| **Previous tests** |  |  |  |
|  | 0 | Ref |  |
|  | 1-2 | 1.009 | (0.61, 1.68) |
| | $\geq 3$ | 0.832 | (0.5, 1.38) |

OR – Odds Ratio.

**Figure S1.**
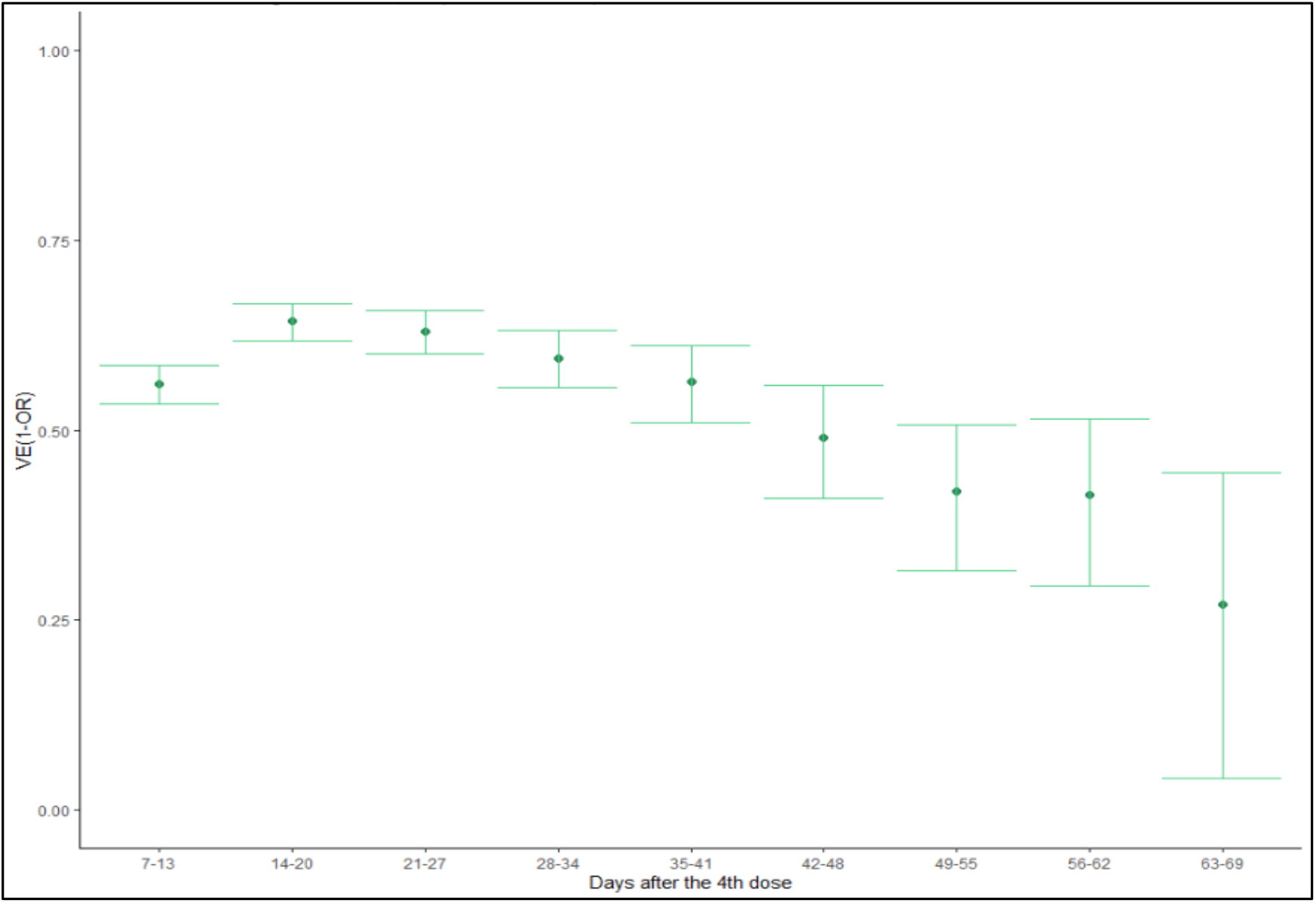
Adjusted fourth dose vaccine effectiveness against SARS-CoV-2 infection relative to three doses. Matched tests approach.

**Figure S2.**
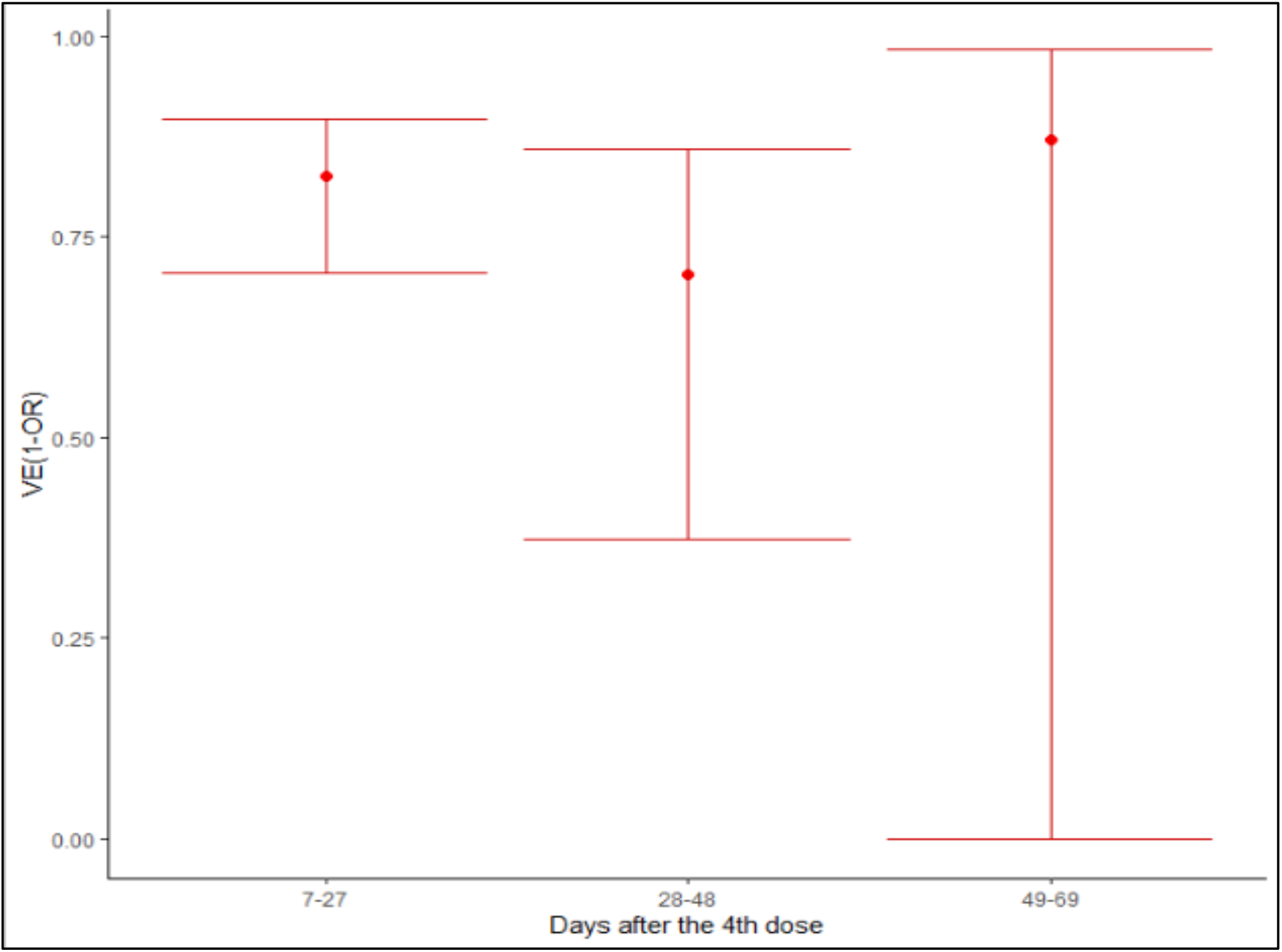
Adjusted fourth dose vaccine effectiveness against SARS-CoV-2 severe disease relative to three doses. Matched tests approach.

## References

1. COVID-19 in Israel dashboard [Internet]. [cited 2022 Mar 5];Available from: https://datadashboard.health.gov.il/COVID-19/general

2. Seow J, Graham C, Merrick B, et al. Longitudinal observation and decline of neutralizing antibody responses in the three months following SARS-CoV-2 infection in humans. Nat Microbiol [Internet] 2020;5(12):1598–607. Available from: https://doi.org/10.1038/s41564-020-00813-8

3. Ruopp MD, Strymish J, Dryjowicz-Burek J, Creedon K, Gupta K. Durability of SARS-CoV-2 IgG Antibody Among Residents in a Long-Term Care Community. J Am Med Dir Assoc [Internet] 2021;22(3):510–1. Available from: https://pubmed.ncbi.nlm.nih.gov/33515497

4. Shrotri M, Navaratnam AMD, Nguyen V, et al. Spike-antibody waning after second dose of BNT162b2 or ChAdOx1. Lancet [Internet] 2021 [cited 2021 Jul 22];0(0). Available from: http://www.thelancet.com/article/S0140673621016421/fulltext

5. Mizrahi B, Lotan R, Kalkstein N, et al. Correlation of SARS-CoV-2-breakthrough infections to time-from-vaccine. Nat Commun 2021;12(1):1–5.

6. Patalon T, Gazit S, Pitzer VE, et al. Odds of Testing Positive for SARS-CoV-2 Following Receipt of 3 vs 2 Doses of the BNT162b2 mRNA Vaccine. JAMA Intern Med [Internet] 2021 [cited 2021 Dec 3];Available from: https://jamanetwork.com/journals/jamainternalmedicine/fullarticle/2786890

7. Bar-On YM, Goldberg Y, Mandel M, et al. Protection of BNT162b2 Vaccine Booster against Covid-19 in Israel. N Engl J Med 2021;385(15):1393–400.

8. Levine-Tiefenbrun M, Yelin I, Alapi H, et al. Viral loads of Delta-variant SARS-CoV-2 breakthrough infections after vaccination and booster with BNT162b2. Nat Med 2021 2021;1–3.

9. SARS-CoV-2 variants in analyzed sequences, Israel [Internet]. [cited 2021 Dec 30];Available from: https://ourworldindata.org/grapher/covid-variants-area?country=~ISR

10. Levine-Tiefenbrun M, Yelin I, Alapi H, et al. Waning of SARS-CoV-2 booster viral-load reduction effectiveness. Nat Commun [Internet] 2022;13(1):1237. Available from: https://doi.org/10.1038/s41467-022-28936-y

11. Patalon T, Saciuk Y, Ma M, et al. Waning Effectiveness of the Third Dose of the BNT162b2 mRNA COVID-19 Vaccine. medRxiv [Internet] 2022 [cited 2022 Feb 27];2022.02.25.22271494. Available from: https://www.medrxiv.org/content/10.1101/2022.02.25.22271494v1

12. Israeli Ministry of Health. Fourth Dose of the Vaccine Approved for People with a Weakened Immune System [Internet]. 2021;Available from: https://www.gov.il/en/departments/news/30122021-05

13. Shalev V, Chodick G, Goren I, Silber H, Kokia E, Heymann AD. The use of an automated patient registry to manage and monitor cardiovascular conditions and related outcomes in a large health organization. Int J Cardiol 2011;152(3):345–9.

14. Weitzman D, Chodick G, Shalev V, Grossman C, Grossman E. Prevalence and factors associated with resistant hypertension in a large health maintenance organization in Israel. Hypertension 2014;64(3):501–7.

15. Chodick G, Heymann AD, Shalev V, Kookia E. The epidemiology of diabetes in a large Israeli HMO. Eur J Epidemiol 2003;18(12):1143–6.

16. Coresh J, Turin TC, Matsushita K, et al. Decline in estimated glomerular filtration rate and subsequent risk of end-stage renal disease and mortality. JAMA - J Am Med Assoc 2014;311(24):2518–31.

17. Dagan N, Barda N, Kepten E, et al. BNT162b2 mRNA Covid-19 Vaccine in a Nationwide Mass Vaccination Setting. N Engl J Med 2021;384(15).

18. Dean NE, Hogan JW, Schnitzer ME. Covid-19 Vaccine Effectiveness and the Test-Negative Design. https://doi.org/101056/NEJMe2113151 [Internet] 2021 [cited 2021 Oct 18];385(15):1431–3. Available from: https://www.nejm.org/doi/full/10.1056/NEJMe2113151

19. Abu-Raddad LJ, Chemaitelly H, Butt AA. Effectiveness of the BNT162b2 Covid-19 Vaccine against the B.1.1.7 and B.1.351 Variants. N Engl J Med [Internet] 2021 [cited 2022 Jan 2];385(2):187–9. Available from: https://www.nejm.org/doi/full/10.1056/NEJMc2104974

20. Chemaitelly H, Tang P, Hasan MR, et al. Waning of BNT162b2 Vaccine Protection against SARS-CoV-2 Infection in Qatar. N Engl J Med 2021;385(24):e83.

21. Goldberg Y, Mandel M, Bar-On YM, et al. Waning Immunity after the BNT162b2 Vaccine in Israel. N Engl J Med 2021;385(24):e85.

22. Ballinger GA. Using Generalized Estimating Equations for Longitudinal Data Analysis: https://doi.org/101177/1094428104263672 2016;7(2):127–50.

23. Chemaitelly H, Ayoub HH, Almukdad S, et al. Duration of protection of BNT162b2 and mRNA-1273 COVID-19 vaccines against symptomatic SARS-CoV-2 Omicron infection in Qatar. medRxiv [Internet] 2022 [cited 2022 Mar 20];2022.02.07.22270568. Available from: https://www.medrxiv.org/content/10.1101/2022.02.07.22270568v1

24. Girard B, Tomassini JE, Deng W, et al. mRNA-1273 Vaccine-elicited Neutralization of SARS-CoV-2 Omicron in Adolescents and Children. medRxiv Prepr Serv Heal Sci [Internet] 2022 [cited 2022 Feb 8];Available from: https://pubmed.ncbi.nlm.nih.gov/35118475/

25. Abu-Raddad LJ, Chemaitelly H, Ayoub HH, et al. Effect of mRNA Vaccine Boosters against SARS-CoV-2 Omicron Infection in Qatar. N Engl J Med [Internet] 2022;Available from: https://doi.org/10.1056/NEJMoa2200797

26. Tenforde MW, Self WH, Naioti EA, et al. Sustained Effectiveness of Pfizer-BioNTech and Moderna Vaccines Against COVID-19 Associated Hospitalizations Among Adults — United States, March–July 2021. Morb Mortal Wkly Rep 2021;70(34):1156.

27. Levine-Tiefenbrun M, Yelin I, Alapi H, et al. Viral loads of Delta-variant SARS-CoV2 breakthrough infections following vaccination and booster with the BNT162b2 vaccine. medRxiv [Internet] 2021 [cited 2021 Oct 5];2021.08.29.21262798. Available from: https://www.medrxiv.org/content/10.1101/2021.08.29.21262798v1

28. Gazit S, Mizrahi B, Kalkstein N, et al. BNT162b2 mRNA Vaccine Effectiveness Given Confirmed Exposure: Analysis of Household Members of COVID-19 Patients. Clin Infect Dis [Internet] 2021 [cited 2021 Dec 3];Available from: https://academic.oup.com/cid/advance-article/doi/10.1093/cid/ciab973/6437962

29. Bar-On YM, Goldberg Y, Mandel M, et al. Protection of BNT162b2 Vaccine Booster against Covid-19 in Israel. N Engl J Med 2021;385(15):1393–400.

30. Mizrahi B, Lotan R, Kalkstein N, et al. Correlation of SARS-CoV-2-breakthrough infections to time-from-vaccine. Nat Commun 2021;12(1):1–5.

